# Cognitive behavioural therapy-based interventions for gastroduodenal disorders of gut-brain interaction: A systematic review

**DOI:** 10.1101/2023.07.20.23292926

**Authors:** Mikaela Law, Isabella Pickering, Esme Bartlett, Gabrielle Sebaratnam, Chris Varghese, Armen Gharibans, Greg O’Grady, Christopher N. Andrews, Stefan Calder

**Affiliations:** The Department of Surgery, The University of Auckland, New Zealand; Alimetry Ltd., Auckland, New Zealand; The Department of Psychological Medicine, The University of Auckland, New Zealand; The Division of Gastroenterology, Cumming School of Medicine, University of Calgary, Canada

**Keywords:** Cognitive behavioural therapy, Functional dyspepsia, Functional gastroduodenal disorders, Functional gastrointestinal disorders, Rumination syndrome, Supragastric belching.

## Abstract

**Objective:** Cognitive behavioural therapy (CBT) is increasingly used to manage Disorders of Gut-Brain Interaction (DGBIs). This systematic review aimed to review the evidence for the effectiveness of CBT-based interventions for patients with gastroduodenal DGBIs.

**Methods:** Medline, Embase, PubMed, Cochrane Central, and Scopus were searched in July 2022. Studies were included if they investigated the effects of a CBT-based intervention on gastrointestinal symptoms and/or psychological outcomes pre- and post-intervention in patients with gastroduodenal DGBIs. Case studies, studies not in English, and studies with patients under 18 years were excluded. Results were synthesised narratively, and standardised effect sizes were calculated where possible.

**Results:** Nine studies (seven RCTs and two pre/post studies) were identified, with data reported in 10 articles (total N=602). The studies investigated patients with functional dyspepsia (n=7), rumination syndrome (n=1), and supragastric belching (n=1). The studies had heterogeneous interventions, methodologies, and outcomes, precluding meta-analysis, as well as a moderate-high risk of bias and high drop-outs rates. Findings demonstrated decreased gastrointestinal symptoms and improved anxiety, depression, and quality of life, from pre- to post-intervention, with medium to large effect sizes for symptoms and small to large effect sizes for psychological outcomes. Efficacy was maintained at follow-up, up to one year later.

**Conclusions:** This review suggests promising evidence that CBT effectively improves gastrointestinal symptoms and psychological outcomes in patients with gastroduodenal DGBIs. However, heterogeneity, risk of bias, and lack of statistical reporting were noted, indicating the need for more robust research and standardisation.

## Introduction

Disorders of gut-brain interaction (DGBIs; formerly known as functional gastrointestinal disorders), where patients have chronic gastrointestinal symptoms, have a staggering 40% global prevalence rate [1,2]. The majority of DGBI research focuses on irritable bowel syndrome (IBS); however, the subset termed gastroduodenal DGBIs remains comparatively under-researched. This group of patients experience chronic symptoms attributed to the gastroduodenal region, such as chronic nausea, vomiting, or belching [3,4]. The ROME IV criteria for gastroduodenal DGBIs include definitions for functional dyspepsia (FD) (including postprandial distress syndrome and epigastric pain syndrome), belching disorders (including excessive supragastric belching (SGB) and excessive gastric belching), nausea and vomiting disorders (including chronic nausea and vomiting syndrome, cyclic vomiting syndrome and cannabinoid hyperemesis syndrome), and rumination syndrome [5]. Although not directly included in the ROME IV subcategories, based on symptomatology, gastroparesis (where chronic nausea and vomiting are associated with delayed gastric emptying) and gastroparesis-like symptoms can also be considered to be closely related to gastroduodenal DGBIs [6,7].

At least 20% of the world’s population meets the criteria for a gastroduodenal DGBI [4] and these disorders are becoming increasingly prevalent over time [8,9]. However, the complex and overlapping symptomology with a lack of organic disease, makes diagnosing and managing these disorders difficult. Consequently, there are limited effective treatment options and reliance on trial-and-error treatment approaches, resulting in high healthcare utilisation and low patient quality of life [1,10–12]. With the increasing prevalence of gastroduodenal DGBIs and their debilitating effects on patients and the healthcare system, there is a need for more effective diagnostic and management options.

Furthermore, dysregulation of the gut-brain axis, a complex bidirectional neurohormonal pathway between the brain and gastrointestinal tract [13–15], may account for the high psychological comorbidity in this patient population [16–19]. Psychological factors, including stress, anxiety, and depression, can worsen gastrointestinal symptoms and quality of life [20–22]. Similarly, the experience of gastrointestinal symptoms can trigger and exacerbate these psychological concerns [23]. Accordingly, due to this bidirectional pathway, a growing body of evidence suggests that psychological interventions can improve mental health and gastrointestinal symptoms in patients with DGBIs [24–26].

Cognitive behavioural therapy (CBT) is one form of psychological intervention that has been well-researched and widely used within gastrointestinal disorders [27]. CBT is an umbrella term for multicomponent psychotherapies focusing on the relationships between thoughts, emotions, behaviours, and physical symptoms [27,28]. CBT-based interventions teach patients to recognise and modify maladaptive thinking styles and/or behaviour patterns that lead to physical symptoms using cognitive and behavioural techniques [29]. By changing these thoughts and behaviours, psychological and physiological symptoms, including gastrointestinal symptoms, can show improvement [30]. Typical components of CBT include psychoeducation, cognitive restructuring, coping skills training, relaxation training, and exposure techniques [27].

Although research has demonstrated that CBT can improve gastrointestinal symptoms, pain, anxiety, depression, and quality of life in patients with gastroduodenal DGBIs, these results have not yet been synthesised or reviewed. Past meta-analyses have shown the beneficial effect of psychological interventions on symptoms and psychological factors in patients with FD [24,25]. Although CBT-based interventions were included in the reviews, these also included other interventions, such as hypnotherapy, psychotherapy, and relaxation therapy, and neither meta-analysis investigated the efficacy of CBT alone. Another meta-analysis found that CBT effectively reduces gastrointestinal symptoms and improves psychology in patients with IBS [31]. However, less is known about its efficacy in gastroduodenal DGBIs. Therefore, this study aimed to systematically review the evidence for the effectiveness of CBT-based interventions for patients with gastroduodenal DGBIs. The primary outcome of interest was the change in patients’ gastrointestinal symptoms from pre- to post-intervention. Secondary outcomes included psychological measures of depression, anxiety, stress, and quality of life.

## Methods

### Protocol

A systematic review protocol was developed based on the Cochrane Handbook for Systematic Reviews of Interventions [32] and the Preferred Reporting Items for Systematic Reviews and Meta-Analyses (PRISMA) statement [33]. The objectives, eligibility criteria, search strategy, risk of bias assessment, and synthesis plan were established before the review and documented in the protocol registered at PROSPERO (ID: CRD42022344902). Deviations from this protocol are detailed in the appropriate sections below.

### Eligibility Criteria

Only primary research studies reported in English were included. Secondary research, unpublished literature, and qualitative research were excluded. No restrictions were placed on the year of publication or study location/setting.

#### Study design

Randomised controlled trials (RCTs) and non-randomised studies with pre- to post-intervention comparisons were included. The protocol was updated to exclude case studies, as these were not considered primary research studies.

#### Participants

Cohorts of patients over 18 years with a diagnosis of a gastroduodenal DGBI (e.g., FD, gastroparesis, supragastric or gastric belching, chronic nausea and vomiting syndrome, cyclic vomiting syndrome, cannabinoid hyperemesis syndrome, or rumination syndrome) were eligible for inclusion. These diagnoses had to be based on a clinician’s diagnosis or by meeting specific diagnostic criteria (e.g. ROME criteria). Studies were excluded if patients had undifferentiated gastrointestinal symptoms or if multiple diagnoses were combined and not separated in the analyses. The a priori protocol encompassed all ages; however, only data from adult patients over 18 years are presented here, as paediatric patients have different diagnostic criteria, disease mechanisms, and treatment approaches.

#### Intervention

For inclusion, patients had to be provided with a CBT-based intervention as the primary intervention of the study. The intervention must include strategies to identify and change maladaptive thinking and behaviour patterns. There were no restrictions on the delivery format or mode of delivery of the intervention.

#### Outcomes

The study had to have measured and reported at least one quantitative measure of gastrointestinal symptoms and/or psychological measures of stress, anxiety, depression, or quality of life at pre and post-intervention.

### Search Strategy

To identify relevant publications for review, the following electronic databases were systematically searched using a standardised search query: Medline, Embase, PubMed, Cochrane Central, and Scopus. The search string combined a set of population and intervention terms within each set with ‘OR’ and between the two sets with ‘AND.’ Search terms related to the outcome measures were not included in the search string as outcome measures are not often well described in abstracts. The outcomes were instead assessed at the screening stage. The final search strategies for two example databases are presented in Table 1.

**Table 1.** Example search strategy syntax for Medline and Embase databases

| Search Strategy Syntax |
| --- |
| 1. (Gastroduodenal disorder or Functional dyspepsia or Non-ulcer dyspepsia or Nonulcer dyspepsia or Dyspepsia or Postprandial distress or Epigastric pain or Gastroparesis or belching disorder or Supragastric belching or Gastric belching or (Chronic nausea and vomiting) or (Nausea and vomiting) or Cyclic vomiting or Cannabinoid hyperemesis or Rumination syndrome or Rumination disorder).mp.<br>2. (CBT or Cognitive behavio* therap* or Cognitive-behavio* therap* or Cognitive behavio* or Cognitive behavio* intervention or Cognitive-behavio* intervention or Cognit* therap* or Behavio* therap* or Mindfulness based cognitive therap* or Mindfulness-based cognitive therap* or MBCT or Cognitive behavio* stress management or CBSM or Dialectical behavio* therap* or Cognitive psychotherap*).mp.<br>3. 1 and 2<br>4. limit 3 to English language |

A search was also conducted by hand of the reference lists of relevant identified articles and the ‘cited by’ feature of Google Scholar was used to search for any other relevant undetected articles. All extracted references from these searches were imported to Covidence (Veritas Health Innovation, Australia) and duplicates were removed. The final search was executed on 27 July 2022. The number of studies identified by the search strategy is shown in Figure 1.

**Figure 1.**
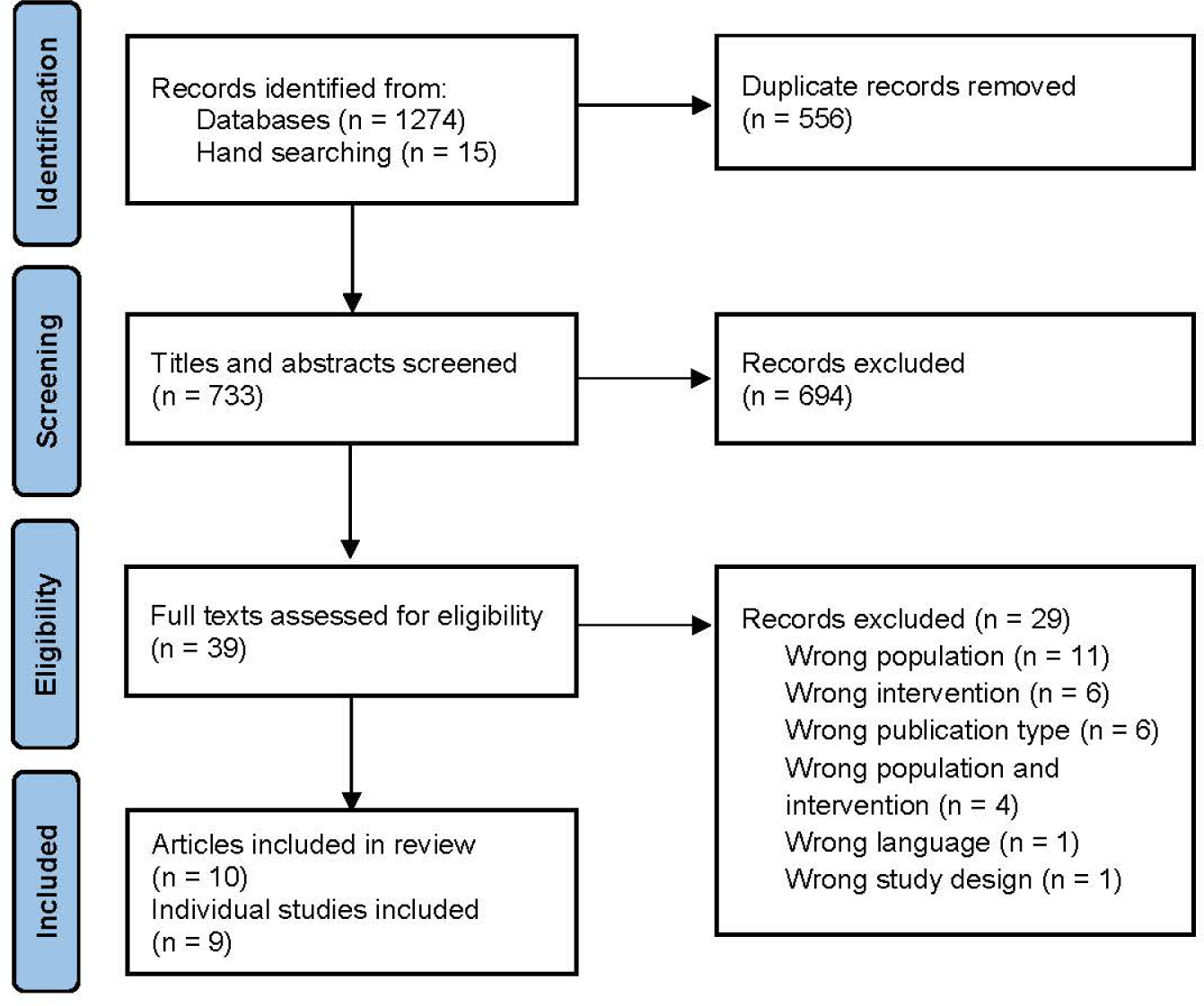
PRISMA flow diagram of the study selection process.

### Screening and Study Selection

Two independent reviewers (ML and GS) screened the studies identified in the search strategy using a two-staged approach using the programme Covidence. In the first screening stage, each reviewer independently screened the titles and abstracts of the identified studies using the eligibility criteria. The article was included in the second stage if a study’s eligibility was uncertain. In the second stage, the two reviewers screened the full texts of the remaining articles to determine the final inclusion or exclusion. Both stages were conducted by the two reviewers independently. The two reviewers’ agreement rate was 98% (κ= 0.78) in the first stage and 95% (κ= 0.86) in the second stage. Disagreements were discussed until an agreement on eligibility was reached. In all instances, an agreement was reached without needing an independent third reviewer. The number of included and excluded studies at each stage of the screening procedure is shown in Figure 1, with reasons for exclusion.

### Data Extraction

Two reviewers (ML and EB) performed data extraction independently into a data extraction template developed per the review’s objectives, using Covidence. The following data were extracted from each included study: publication details (i.e. title, authors, year of publication, country of origin, source of funding), study design, sample characteristics (sample size, age, gender split, diagnosis, diagnostic criteria used), intervention details (type of interventions, delivery method, format, practitioner, number of sessions, duration), and outcome measures (symptom and/or psychological measures used, timepoints collected).

Key findings were extracted using two methods to assess two different dimensions of efficacy: (1) as within-group pre-post outcome comparisons in the CBT group, and (2) as between-group differences in the outcomes at post-intervention between the CBT group and the other group(s). Data was only extracted for relevant outcome measures relating to gastrointestinal symptoms and/or psychological outcomes. Where possible, data was extracted as an intention-to-treat using all available follow-up data. Missing data was reported as not stated (NS). Data were extracted independently by the two reviewers, with an agreement rate of 94%. Inconsistencies were discussed, with final decisions made by a third independent reviewer (GS).

### Risk of Bias

The studies were assessed independently by two reviewers (ML and EB) for risk of bias using the Cochrane RoB 2 tool [34] for the RCTs and the ROBINS-I tool [35] for pre/post-cohort studies. There was an 81% agreement between the two reviewers for the ROB 2 tool and 96% for the ROBINS-I tool. Conflicts were discussed, with a third independent reviewer making the final decisions (GS).

## Data Analysis

The studies had heterogeneous interventions, methodologies, and outcome measures, in addition to inconsistent statistical reporting. Therefore, the results could not be meaningfully combined for a meta-analysis. The data extracted from the studies were thus narratively synthesised. The study results were also converted to standardised mean difference effect sizes (Cohen’s *d*), where data was available, to make the main effects more comparable. Cohen’s *d*s were calculated for both within-group and between-group effects using the standard formula;

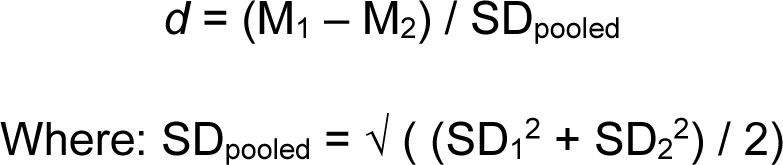

Within-group effect sizes were calculated using the standardised mean difference in the outcomes of interest between the pre- and post-intervention and pre-intervention and follow-up timepoints in the CBT group. Between-group effect sizes were calculated using the standardised mean difference in the outcomes of interest between the CBT and comparator group(s) at the post-intervention and follow-up timepoints. All effect sizes were calculated using intention-to-treat data where available. However, not all studies reported the means and standard deviations required to calculate these effect sizes. In these studies, effect sizes were reported as having no information available (NI) and results were instead narratively summarised in-text.

## Results

### Search Results

The search strategy resulted in 1289 articles, which were screened for eligibility. Following the initial title and abstract screening, 39 full-text articles were screened. This screening resulted in nine individual studies, with data reported in 10 articles included in this review (including a total of 602 participants). Two articles were merged into one study, per Cochrane guidelines, as they reported on the same study’s results; Glasinovic et al. [36] reported the initial results, while Sawada et al. [37] reported the extended analyses and follow-up data. A summary PRISMA diagram of the screening process is displayed in Figure 1.

#### Near misses

Four articles were considered “near misses” for this review (studies that did not meet the eligibility criteria but were very close to being included). Dast et al. [38] was excluded because although their intervention incorporated CBT-based components, it was primarily focused on mindfulness-based techniques. Punkkinen et al.’s [39] behavioural therapy for SGB included only behavioural techniques (i.e. education and diaphragmatic breathing) but did not include cognitive techniques. Niesen et al. [40] included a nurse-led CBT approach for adults with functional abdominal pain; however, the results for patients with FD were combined with patients with functional abdominal pain, so the data for FD could not be extracted separately.

### Study Characteristics

All nine studies were primary studies published as journal articles. The key characteristics of the nine included studies are presented in Table 2. The publication dates ranged from 1994 to 2021. Four studies were carried out in Europe [36,37,41–43], three in Iran [44–46], one in the United States of America [47], and one in Singapore [48]. Seven studies were RCTs with comparator groups [41–46,48]. These comparator groups included standard control groups, as well as other comparative interventions, including; intensive medical care (+/ progressive muscle relaxation) [43] and pharmacological interventions [44,46]. The remaining two were pre/post-cohort studies, with no comparator groups [36,37,47].

#### Sample characteristics

The study sample sizes ranged from 10 to 158. However, most studies had a high number of dropouts, with 40% of the patients dropping out across all nine studies. The majority of the studies (n=7) investigated patients with FD, while the remaining two studies investigated patients with rumination syndrome [47] and SGB [36,37]. Most studies (n=7) used the Rome symptomology criteria to diagnose patients, with five of these studies also using negative results from other medical tests, such as endoscopies, to confirm this diagnosis [41,44–46,48]. Murray et al. [47] used the Pica, ARFID, Rumination, Disorder Interview (PARDI) as a confirmatory assessment. On the other hand, Haug et al. [42] used a clinician’s diagnosis of FD without a named diagnostic criterion. The last study used 24-hour MII pH monitoring to measure the amount of supragastric belches a patient had over 24 hours to confirm the diagnosis of SGB [36,37]. No studies compared patients to healthy controls.

#### Intervention details

The interventions consisted of a variety of CBT-based techniques. Three studies used a traditional CBT intervention [36,37,43,47] and another two implemented cognitive psychotherapy [41,42]. The remaining studies employed other therapies that utilise CBT techniques, including metacognitive therapy (MCT) [44], cognitive behavioural stress management (CBSM) [45], mindfulness-based cognitive therapy (MBCT) [48], and dialectical behaviour therapy (DBT) [46]. All interventions were conducted in person, with no digital interventions included in the review.

One study reported that the intervention was conducted in a group format [48], and three studies did not specifically state how the intervention was delivered [44–46]. The remaining studies were one-on-one with the intervention practitioner. However, the delivery method of the intervention was not always explicitly stated in the manuscripts and instead assumed based on the intervention details. Generally, the CBT-based interventions were run by psychologists, therapists, or trained instructors [44–46]. The frequency of CBT sessions ranged from 4-20 sessions, and the duration of each session varied from 45 minutes to two hours. One study also included a half-day retreat as part of the programme [48].

#### Outcome measures

Table 2 shows the various outcome measures for gastrointestinal symptoms and psychological variables used across the studies. Two studies only measured gastrointestinal symptoms [45,47], two studies only measured psychological outcomes [44,48], and the remaining five measured both [36,37,41–43,46]. All studies measured the outcomes before and after the intervention. Seven also had a follow-up timepoint, which ranged from 1 to 12-months after the end of the intervention.

As shown in Table 2, all studies that measured gastrointestinal symptoms included at least one self-reported symptom scale, with no two studies using the same symptom measures. Alongside these self-reported scales, Murray et al. [47] also used the PARDI interview to gather clinician-reported symptoms and the study by Glasinovic et al. [36], and Sawada et al. [37] used 24-hour MII pH monitoring to objectively measure the number of belches and reflux episodes. All psychological outcomes were measured using self-report questionnaires and included measures of depression [41–44,48], anxiety [41–44,46,48], stress [48], and health-related quality of life [36,37,43,48].

### Risk of Bias

Figure 2 presents the risk of bias scores from the RoB 2 tool for the seven RCTs. Overall, all the RCTs had concerns or a high risk of bias. Bias concerns were raised due to a lack of reporting about randomisation, whether there were deviations in the intended interventions, and whether the data reported was in accordance with a pre-specified analysis plan. Many studies also had incomplete outcome data due to high dropout numbers. Due to the nature of the interventions, it was impossible to blind participants and practitioners to group allocation, which may have biased the outcome measurement in all studies.

**Figure 2.**
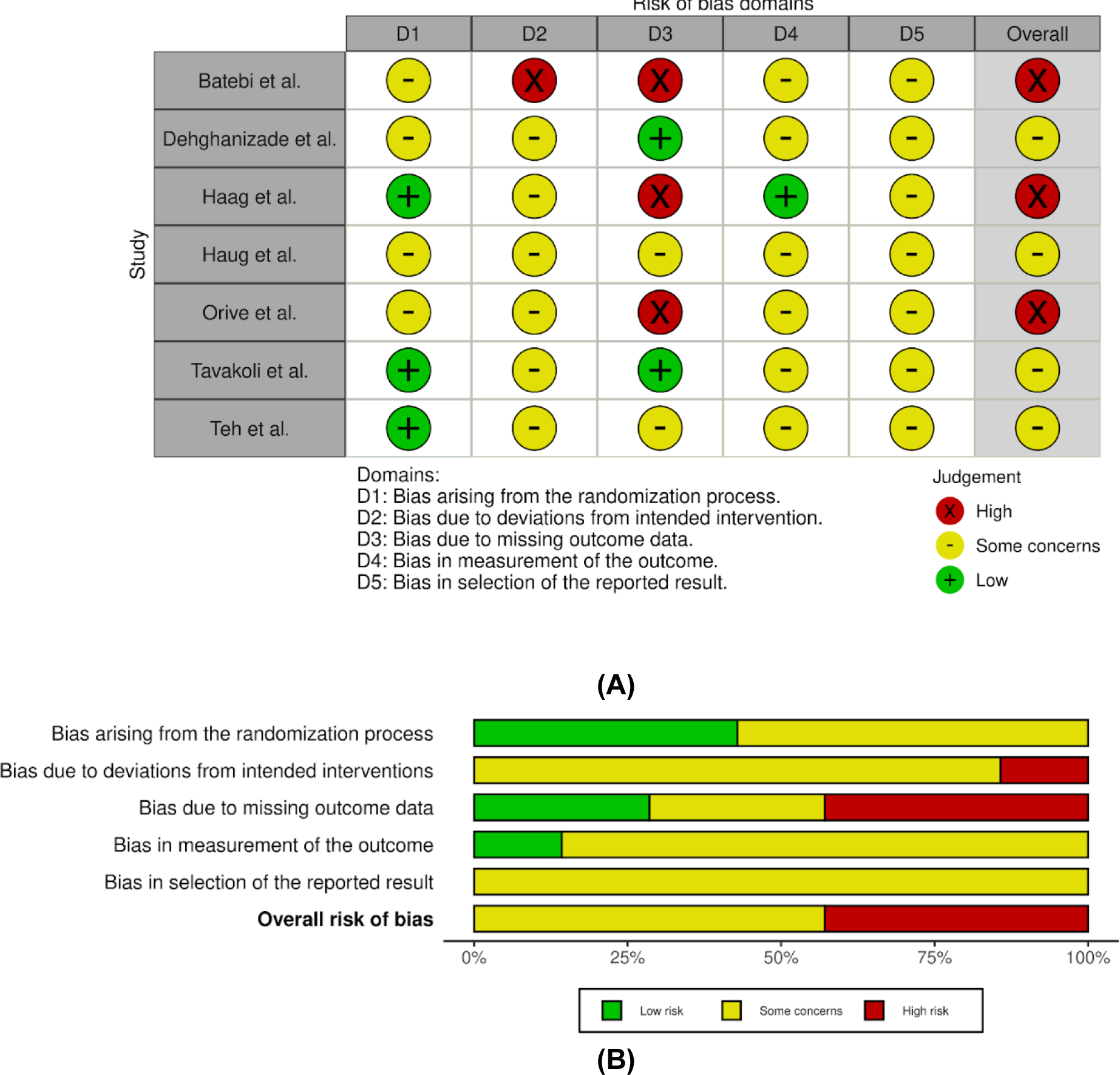
Risk of bias summary from the ROB 2 tool. (A) Summary of risk of bias for each trial. (B) The risk of bias graph about each domain is presented as a percentage across all included studies. Figures were generated using robvis [49].

Figure 3 presents the risk of bias scores from the ROBINS-I tool for the two pre/post-cohort studies. Both studies showed a moderate risk of bias arising from missing outcome data and high dropout rates. Bias in the measurement of outcomes could not be assessed in either study, as these studies only had one intervention group.

**Figure 3.**
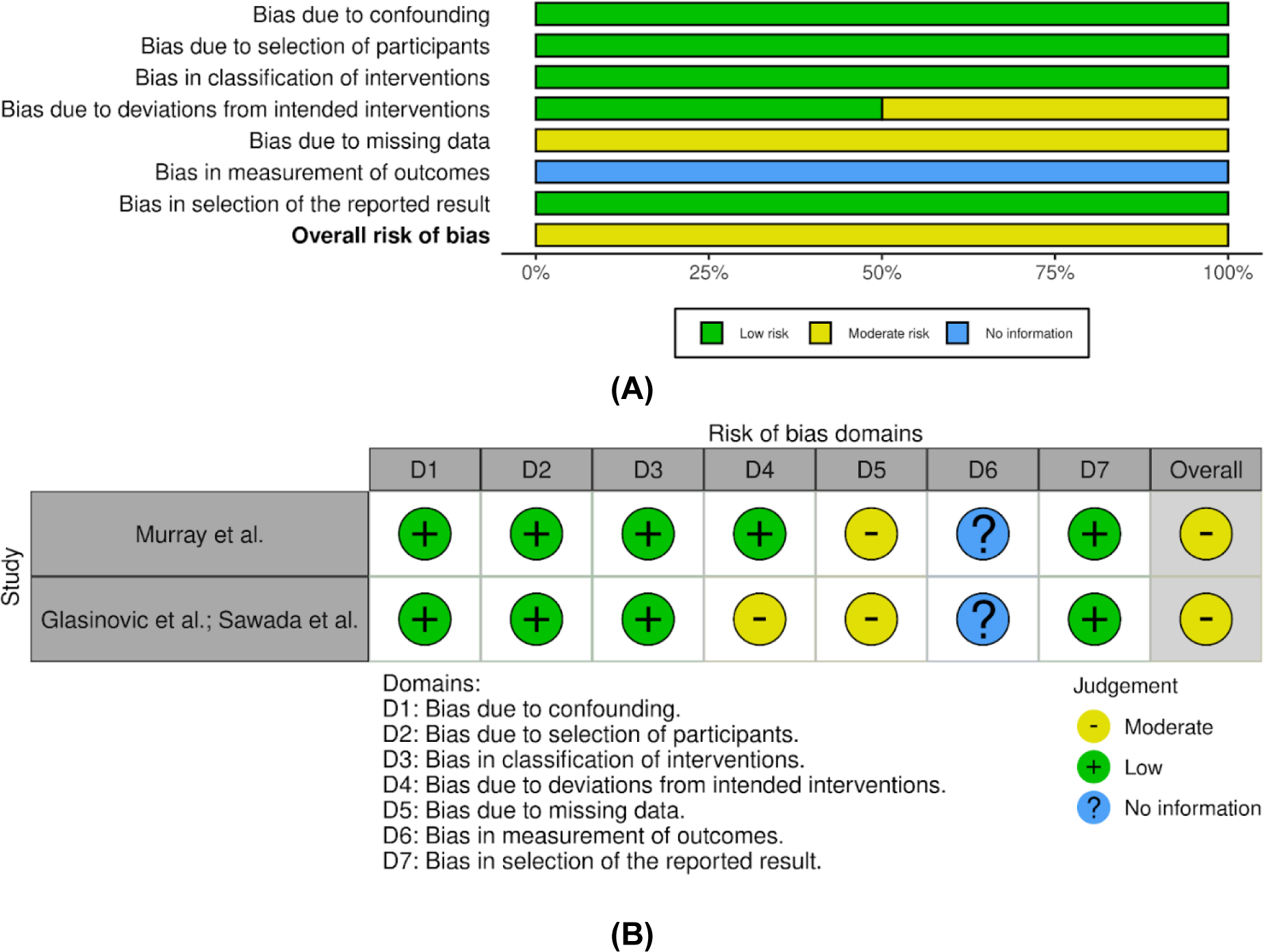
Risk of bias summary from the ROBINS-I tool. (A) Summary of risk of bias for each trial. (B) The risk of bias graph about each domain is presented as a percentage across all included studies. Figures were generated using robvis [49].

### Summary of Findings

Key results from the studies are shown in Table 3 for within-group effects and Table 4 for between-group effects (CBT vs comparator groups) and are synthesised descriptively below.

**Table 3.** *Characteristics of studies included in the review*

| Study | Country | Funding | Study design | Patient diagnosis (diagnostic criteria) | Age, M (range, if stated) | % female | Study groups (sample size) | CBT intervention delivery | CBT intervention duration | GI symptom outcomes | Psychological outcomes | Follow-up duration |
| --- | --- | --- | --- | --- | --- | --- | --- | --- | --- | --- | --- | --- |
| Batebi et al., 2020 [44] | Iran | NS | RCT | Functional Dyspepsia (Rome IV) | 37 (20-50) | 55% | MCT (N=20, 3 dropouts)<br>Nortriptyline (N=20, 4 dropouts)<br>SMT (N=25, 10 dropouts) | NS | 10x, 45-minute sessions over 10 weeks |  | Anxiety (HAM-A)<br>Depression (HDRS) | 3 months |
| Dehghanizade et al., 2015 [45] | Iran | NS | RCT | Functional Dyspepsia (Rome III) | 29 | NS | CBSM (N=15)<br>No intervention (N=15) | NS | 10x sessions (length of sessions and programme duration NS) | NDI (symptom checklist) |  | 1 month |
| Glasinovic et al., 2018 [36];<br>Sawada et al., 2019 [37] | United Kingdom | University | Pre/Post Cohort | Supragastric Belching (MII-pH monitoring) | 45 (20-72) | 51% | CBT (N=51, 12 dropouts) | Individual, face-to-face, with a therapist | 4x sessions over 10 weeks (length of sessions NS) | Belching Symptom Severity VAS<br>Number of belches and reflux episodes (via MII-pH monitoring) | HRQoL (SF-36) | 6-12 months |
| Haag et al., 2007 [43] | Germany | Govt | RCT | Functional Dyspepsia (Rome II) | 45 (35-54) | 64% | Intensive medical therapy + CBT (N=28, 7 dropouts)<br>Intensive medical therapy + PMR (N=20, 10 dropouts)<br>Intensive medical therapy (N=28, 8 dropouts)<br>SMT (N=24, 19 dropouts) | Individual, face-to-face, with a psychologist | 20x, 1-hour sessions over 20 weeks | The Symptom Intensity and Severity Scale | HRQoL (SF-36)<br>Anxiety and Depression (HADS) | 12 months |
| Haug et al., 1994 [42] | Norway | NS | RCT | Functional Dyspepsia (Clinician diagnosis) | 40 | 59% | Cognitive psychotherapy (N=50, 7 dropouts)<br>No treatment (N=50, 5 dropouts) | Individual, face-to-face, with a psychiatrist | 10x 50-minute sessions over 4 months | Changes in dyspeptic symptoms | Anxiety (STAI-I-II)<br>Depression (BDI) | 12 months |
| Murray et al., 2021 [47] | United States of America | Private/charity | Pre/Post Cohort | Rumination Syndrome (Rome IV) | 36 (20-67) | 50% | CBT (N=10, 2 dropouts) | Individual, face-to-face, with a psychologist | 5-8x, 45-minute sessions over 5 weeks | Self-monitored daily regurgitation frequency<br>PARDI |  | 3 months |
| Orive et al., 2015 [41] | Spain | Govt | RCT | Functional Dyspepsia (Rome III) | 46 | 82% | Psychotherapy (N=76, 18 dropouts)<br>SMT (N=82, 12 dropouts) | Individual, face-to-face, with a therapist | 10x, 50-minute sessions over 10 weeks | GDSS<br>DRHS<sup>a</sup> | Anxiety and Depression (HADS) | 6 months |
| Tavakoli et al., 2020 [46] | Iran | None | RCT | Functional Dyspepsia (Rome III) | 28 | 68% | DBT (N=20)<br>Anxiolytic (N=20)<br>No intervention (N=20) | NS | 8x, 90-minute sessions over 8 weeks | PAGI-SYM | Anxiety (BAI) | NA |
| Teh et al., 2021 [48] | Singapore | Hospital | RCT | Functional Dyspepsia (Rome III) | 49 | 46% | MBCT (N=15, 3 dropouts)<br>SMT (N=13, 2 dropouts) | Group, face-to-face, with a mindfulness instructor | 8x, 2-hour sessions (+ a half-day retreat) over 8 weeks |  | Depression, Anxiety and Stress (DASS-21)<br>HRQoL (SF-NDI and EuroQoL-VAS) | NA |

Notes: NS= not stated, RCT= randomised controlled trial, CBT= cognitive behavioural therapy, MCT= metacognitive therapy, SMT= standard medical treatment, CBSM= cognitive stress behavioural management, PMR= progressive muscle relaxation, DBT= dialectical behaviour therapy, MBCT= mindfulness-based cognitive therapy, NDI= Nepean Dyspepsia Index, VAS= visual analogue scale, PARDI= Pica, AFRID and Rumination Disorder Interview, GDSS= The Glasgow Dyspepsia Severity Score, DRHS= The Dyspepsia Related Health Scale, PAGI-SYM= the patient assessment of upper gastrointestinal symptom severity index, HAM-A= Hamilton Anxiety Rating Scale, HDRS= Hamilton Depression Rating Scale, HRQoL= health-related quality of life, SF-36= 36-Item Short Form Survey, HADS= Hospital Anxiety and Depression Scale, STAI= State-Trait Anxiety Inventory, BDI= Beck Depression Inventory, BAI= Beck Anxiety Inventory, DASS= Depression Anxiety and Stress Scale, <sup>a</sup>Only Severity of Common Symptoms Subscale.

**Table 3.** Within-group, pre- to post-intervention effects, reported for the cognitive behavioural therapy group of each included study

| Study | Gastrointestinal symptoms |  |  |  | Psychological outcomes |  |  |  |
| --- | --- | --- | --- | --- | --- | --- | --- | --- |
|  | Outcomes | Direction of effect | Pre-post effect sizes (*d*) | Pre-follow-up effect sizes (*d*) | Outcomes | Direction of effect | Pre-post effect sizes (*d*) | Pre-follow-up effect sizes (*d*) |
| Batebi et al., 2020 [44] |  |  |  |  | a. Anxiety<br>b. Depression | a. ↓<br>b. ↓ | a. 2.36<br>b. 2.30 | a. 2.51<br>b. 2.49 |
| Dehghanizade et al., 2015 [45] | a. Symptom severity | a. ↓ | a. 2.22 | a. 2.85 |  |  |  |  |
| Glasinovic et al., 2018 [36]; Sawada et al., 2019 [37] | a. Symptom severity<br>b. Supragastric belches<br>c. Gastric belches<br>d. Acid and non-acid reflux episodes<br>e. Acid reflux episodes | a. ↓<br>b. ↓<br>c. ↑<br>d. ↓<br>e. ↓ | a. 1.39<br>b. 0.58<br>c. 0.28<br>d. 0.41<br>e. 0.59 | a. 1.42 | a. Physical function<br>b. Role limitation physical<br>c. Bodily pain<br>d. General health<br>e. Vitality<br>f. Social function<br>g. Role limitation emotional<br>h. Mental health | a. ↑<br>b. ↑<br>c. ↑<br>d. ↑<br>e. ↑<br>f. ↑<br>g. ↑<br>h. ↑ | a. 0.29<br>b. 0.24<br>c. 0.36<br>d. 0.38<br>e. 0.40<br>f. 0.50<br>g. 0.18<br>h. 0.30 |  |
| Haag et al., 2007 [43] | a. Symptom intensity | a. ↓ | NI | NI | a. Mental HRQoL<br>b. Physical HRQoL<br>c. Total HRQoL<br>d. Anxiety<br>e. Depression | a. ↑<br>b. ↑<br>c. ↑<br>d. ↓<br>e. ↓ | NI | NI |
| Haug et al., 1994 [42] | a. Epigastric pain<br>b. Heartburn<br>c. Nausea<br>d. Bloating<br>e. Lower abdominal pain<br>f. Diarrhoea<br>g. Constipation<br>h. Days of epigastric discomfort | a. ↓<br>b. ↓<br>c. ↓<br>d. ↓<br>e. ↓<br>f. ↓<br>g. ↓<br>h. ↓ | NI | NI | a. State anxiety<br>b. Trait anxiety<br>c. Depression | a. ↓<br>b. ↓<br>c. ↓ | NI | NI |
| Murray et al., 2021 [47] | a. Regurgitation frequency (self-report)<br>b. Regurgitation frequency (interview) | a. ↓<br>b. ↓ | NI | NI |  |  |  |  |
| Orive et al., 2015 [41] | a. Symptom Severity (GDSS)<br>b. Symptom Severity (DRHS subscale) | a. ↓<br>b. ↑<sup>a</sup> | a. 1.67<br>b. 0.70 | a. 1.64<br>b. 0.54 | a. Anxiety<br>b. Depression | a. ↓<br>b. ↓ | a. 0.20<br>b. 0.45 | a. 0.45<br>b. 0.35 |
| Tavakoli et al., 2020 [46] | a. Symptom Severity | a. ↓ | a. 1.42 |  | a. Anxiety | a. ↓ | a. 0.99 |  |
| Teh et al., 2021 [48] |  |  |  |  | a. DASS Total<br>b. Depression<br>c. Anxiety<br>d. Stress<br>e. HRQoL (SF-NDI)<br>f. HRQoL (EuroQoL) | a. ↓<br>b. ↓<br>c. ↓<br>d. ↓<br>e. ↑<br>f. ↑ | NI |  |

*Note:* ↓= Scores decreased overtime; ↑= Scores increased overtime; NI= publication did not have enough information to calculate the within-group effect sizes; HrQoL= Health-related quality of life; GDSS= The Glasgow Dyspepsia Severity Score; DRHS= The Dyspepsia Related Health Scale; DASS= Depression, Anxiety and Stress Scale; SF-NDI= Short Form Nepean Dyspepsia Index.
<sup>a</sup>An increase in this subscale represents improved symptoms.

**Table 4.** Between-group effect sizes for the cognitive behavioural therapy group vs. other comparator groups at the post-intervention and follow-up timepoints of each randomised controlled trial

| Study | Comparator group | Gastrointestinal symptoms |  |  |  | Psychological outcomes |  |  |  |
| --- | --- | --- | --- | --- | --- | --- | --- | --- | --- |
|  |  | Outcomes | Direction of effect | Effect size (*d*) for differences at post-intervention | Effect size (*d*) for differences at follow-up | Outcomes | Direction of effect | Effect size (*d*) for differences at post-intervention | Effect size (*d*) for differences at follow-up |
| Batebi et al., 2020 [44] | Nortriptyline |  |  |  |  | a. Anxiety | a. ↓ | a. 1.39 | a. 1.75 |
|  |  |  |  |  |  | b. Depression | b. ↓ | b. 1.02 | b. 1.53 |
|  | Standard medical treatment |  |  |  |  | a. Anxiety | a. ↓ | a. 3.49 | a. 3.73 |
|  |  |  |  |  |  | b. Depression | b. ↓ | b. 3.64 | b. 4.00 |
| Dehghanizade et al., 2015 [45] | No intervention | a. Symptom severity | a. ↓ | a. 3.74 | a. 5.55 |  |  |  |  |
| Haug et al., 1994 [42] | No intervention | a. Epigastric pain | a. ↓ | NI | NI | a. State anxiety | a. ↓ | NI | NI |
|  |  | b. Heartburn | b. - |  |  | b. Trait anxiety | b. ↑ |  |  |
|  |  | c. Nausea | c. ↓ |  |  | c. Depression | c. ↓ |  |  |
|  |  | d. Bloating | d. - |  |  |  |  |  |  |
|  |  | e. Lower abdominal pain | e. - |  |  |  |  |  |  |
|  |  | f. Diarrhea | f. ↓ |  |  |  |  |  |  |
|  |  | g. Constipation | g. - |  |  |  |  |  |  |
|  |  | h. Days of epigastric discomfort | h. ↓ |  |  |  |  |  |  |
| Orive et al., 2015 [41] | Standard medical treatment | a. Symptom Severity (GDSS) | a. ↓ | a. 0.53 | a. 0.29 | a. Anxiety | a. ↓ | a. 0.04 | a. 0.34 |
|  |  | b. Symptom Severity (DRHS subscale) | b. ↑<sup>a</sup> | b. 0.49 | b. 0.50 | b. Depression | b. ↓ | b. 0.29 | b. 0.32 |
| Tavakoli et al., 2020 [46] | Anxiolytic drug | a. Symptom Severity | a. ↓ | a. 1.27 |  | a. Anxiety | a. ↑ | a. 0.21 |  |
|  | No intervention | a. Symptom Severity | a. ↓ | a. 0.40 |  | a. Anxiety | a. ↓ | a. 0.27 |  |

Note: ↓ = Scores were lower in the CBT group than the comparator group post-intervention; ↑ = Scores were higher in the CBT group than the comparator group at post intervention; - = Scores were the same between CBT group and the comparator group post-intervention; NI = publication did not have enough information to calculate the between-group effect sizes; HrQoL = Health-related quality of life; GDSS = The Glasgow Dyspepsia Severity Score; DRHS = The Dyspepsia Related Health Scale; DASS = Depression, Anxiety and Stress Scale; SF-NDI = Short Form Nepean Dyspepsia Index.
<sup>a</sup>An increase in this subscale represents improved symptoms.
Haag et al. and Teh et al. only reported change scores, and therefore differences between the groups at the post-intervention could not be reported.

#### Gastrointestinal symptom severity

Six studies [36,37,41–43,45,46] assessed pre-to-post differences in symptom severity, all of which found decreases at the post-CBT timepoint.

Among studies where effect sizes could be calculated, Cohen’s *d* ranged from 0.70 to 2.22 (*M*=1.48), indicating large effect sizes. Five studies [36,37,41–43,45] also assessed symptom severity at follow-up, showing the large decreases in symptom severity were maintained at 1-month [45], 6-months [41], and 12-months [36,37,42,43] post-intervention, with Cohen’s *d* ranging from 0.54 to 2.85 (*M*=1.61).

Four studies [41,42,45,46] also tested the effectiveness of CBT on symptom severity against comparator groups, all finding that symptom severity was lower in the CBT group post-intervention compared with the comparator groups. Cohen’s *d* ranged between 0.40 and 3.74 (*M*=1.29), indicating large effect sizes. Three studies [41,42,45] showed that these effects were maintained long-term, with Cohen’s *d* ranging between 0.29 and 5.55 (*M*=2.92).

#### Gastrointestinal symptom episodes

Three studies [36,37,42,47] assessed pre-to-post differences in the frequency of gastrointestinal symptom episodes, finding decreases in reflux episodes, supragastric belches [36,37], regurgitation frequency [47], and days of epigastric discomfort [42]. Where available, Cohen’s *d* ranged from 0.41 to 0.59 (*M*=0.52), indicating medium effect sizes. However, one study [36,37] found that gastric belches increased in the CBT group from pre- to post-intervention, despite all other symptom episodes decreasing over time in this study. All three studies assessed differences in symptom episode frequency at a follow-up timepoint, finding that decreases were maintained at 3-months [47] and 12-months [36,37,43]. One study [42] compared days of epigastric discomfort between the CBT group and a no-intervention group, with the CBT group having fewer days of epigastric discomfort post-intervention, with the effect maintained at 12-months.

#### Depression

All five studies [41–44,48] that measured depression showed a decrease from pre- to post-intervention in the CBT group. Cohen’s *d* ranged from 0.30 to 0.45 (*M*=0.37), indicating small effect sizes, which were maintained at 3-months [44], 6-months [41], and 12-months [42,43]. Three studies [41,42,44] also found that depression was lower in the CBT group post-intervention, compared to the comparator group(s), with Cohen’s *d* ranging between 0.29-3.64 (*M*=2.48), indicating variability, with an average large effect size. Follow-up was also assessed in these three studies, with the differences between the CBT and comparator groups increasing over time, with available effect sizes varying from *d*=0.32-4.00 (*M*=2.93) [41,44].

#### Anxiety

All six studies [41–44,46,48] that assessed pre-to-post differences in anxiety found a decrease over time in the CBT group. Cohen’s *d* ranged from 0.20 to 2.36 (*M*=1.83), indicating variability in effect sizes. Four studies [41–44] found that anxiety remained lower than pre-intervention at 3-months [44], 6-months [41], and 12-months [42,43] post-intervention. Four studies also compared anxiety between CBT and a comparator group at the post-intervention timepoint, showing mixed effects. The studies reported significantly lower anxiety post-intervention in the CBT group compared to nortriptyline [44], standard medical treatment [41,44], and no intervention [42,46], with Cohen’s *d* ranging widely between 0.04 and 3.49 (*M*=1.77), with an average large effect size. However, Tavakoli et al. [46] found that despite having lower anxiety post-intervention than the no-intervention group, the CBT group had higher anxiety than the group that received anxiolytic medication. Haug et al. [42] also found that the CBT group had higher trait anxiety scores post-intervention than the group that received no intervention.

#### Stress

Only one study [48] assessed pre-to-post differences in stress, with findings showing that the CBT group decreased stress from pre-to post-intervention. No follow-up was assessed in this study.

#### Health-related quality of life

All three studies [36,37,43,48] that measured health-related quality of life showed an increase in the CBT group from pre-to post-intervention, with one study finding that these effects were maintained at 12-months [43].

## Discussion

This systematic review aimed to review the available evidence on the effectiveness of CBT-based interventions for patients with gastroduodenal DGBIs. Nine studies, with data from 10 articles, were included in the review. Seven studies were published in the last 10 years, demonstrating that research into the psychological management of gastroduodenal DGBIs is growing.

Overall, the findings from the review show promising evidence supporting the effectiveness of CBT-based interventions for improving both gastrointestinal symptoms and psychological outcomes in patients with gastroduodenal DGBIs. These effects were generally maintained at follow-up, up to 12 months after the end of the intervention. Results included consistent reductions in gastrointestinal symptom severity and symptom episodes, including reflux episodes, regurgitation frequency, days of epigastric discomfort, and the number of supragastric belches. However, CBT did not appear to have a benefit on gastric belching in the one study that measured this. The effect of CBT on psychological outcomes was less consistent but still indicated promising evidence. Overall, patients who received CBT-based interventions experienced improvements in depression, state anxiety, stress, and health-related quality of life, albeit with smaller effect sizes than gastrointestinal symptoms.

However, the efficacy of CBT compared to neuromodulating pharmacological therapies was mixed. While one study showed that CBT was less efficacious for anxiety reduction than anxiolytics [46], another study demonstrated that CBT was more effective at reducing anxiety and depression than antidepressants [44]. Regardless, these two studies found that CBT and pharmacological interventions were more effective than standard medical treatment for gastroduodenal disorders [44,46]. These findings suggest the potential utility of CBT to enhance existing pharmacological treatment or as a standalone treatment; however, more research is needed to explore this further.

The findings from this review are consistent with meta-analyses across the gastrointestinal tract, which have found that psychological interventions generally improve gastrointestinal symptoms and psychological outcomes in patients with DGBIs [24,25]. The current review expands on this evidence by demonstrating that these effects are specific to CBT-based interventions, particularly for patients with gastroduodenal DBGIs. CBT-based interventions have also been efficacious for patients with other DGBIs, such as IBS, where a large evidence base already exists [31,50–52]. However, further research is needed to translate IBS-specific treatment protocols and guidelines to gastroduodenal DGBIs.

The encouraging findings from this review suggest that CBT-based interventions should be considered another potential treatment avenue for gastroduodenal DGBIs. Clinical guidelines for these conditions recommend psychological interventions as a potential treatment avenue [26,53] and patients have reported openness to psychological interventions as part of their management [54]. CBT-based interventions are low-risk and safe compared to traditional pharmacotherapy or surgical treatments and may particularly benefit patients with psychological comorbidities or strong gut-brain influences on their symptoms. Recent developments in gastric physiology testing using body surface gastric mapping with simultaneous symptom tracking may be able to distinguish these patients from those with neuromuscular dysfunction [55].

Therefore, CBT-based interventions could be incorporated as part of a multidisciplinary management plan for patients with gastroduodenal DGBIs, which can be tailored to the patient’s individual needs and involve the collaboration of gastroenterologists and psychologists. Research into the effectiveness of multidisciplinary integrated treatment approaches has shown improvements in symptom management and psychological outcomes compared to standard medical treatment [56–59]. However, there are considerable barriers to in-person CBT, including patient stigma, cost, and accessibility. Digital CBT-based interventions could help remove these barriers and increase accessibility; however, none of the studies included in this review assessed the effects of digital CBT. IBS research has previously demonstrated the effectiveness and acceptability of CBT-based interventions delivered via the Internet and mobile applications [60–62], with similar efficacy to in-person therapy [63,64]. Therefore, it is likely that patients with gastroduodenal DGBIs would similarly benefit from using digital CBT through web browsers or mobile apps.

### Overall Completeness and Applicability of Evidence

Despite this promising evidence, definitive conclusions are difficult to make from these studies due to heterogeneous methodologies, comparators, outcomes, and interventions. Although all studies utilised CBT-based interventions that incorporated strategies to identify and change maladaptive thinking and behaviour patterns, a variety of techniques were used. Further research is needed to identify which components of CBT are the most efficacious.

Furthermore, all RCTs included involved patients with FD, whilst the studies for SGB and rumination syndrome only include pre/post cohort designs. Other gastroduodenal DGBIs, such as chronic nausea and vomiting syndrome or cyclic vomiting syndrome, did not have any available evidence, despite being common and having a strong biological rationale for using CBT. Therefore, the majority of the evidence from this review applies to patients with FD and may not necessarily extrapolate to patients with other gastroduodenal DGBIs. The current review also excluded studies where patients had undifferentiated DGBIs or those with multiple diagnoses combined, which may have excluded potentially important data.

### Quality of the Evidence

The included studies showed a moderate to high risk of bias, which may have affected the quality of evidence in the review. This risk of bias was primarily due to missing data due to inconsistent statistical reporting and high numbers of dropouts. Patients who dropped out will likely have different outcomes than those who did not. Some studies used an intention-to-treat analysis to account for these differences; however, most studies did not statistically account for missing data. We utilised standardised effect sizes in an attempt to standardise the data; however, a lack of statistical reporting meant that these effect sizes could not be calculated for all studies. Therefore, direct comparisons between all studies were not possible, limiting the statistical power of this review.

Lastly, a lack of blinding was inevitably identified as a risk of bias in all studies, which may have led to potential placebo effects and observation biases. Previous reviews have suggested that active psychotherapy control groups be used to counter this issue and to account for the effect of supportive care and communication in psychological intervention studies [24]. Based on these bias assessments, future research should focus on increasing the methodological quality of the studies and using more rigorous reporting standards.

### Potential Biases in the Review Process

Agreement between the two independent reviewers was high for both eligibility screening and data extraction, indicating that the review had a rigorous and clear methodology. We also did not limit the review to only RCTs, which allowed the inclusion of other relevant study designs. However, this review is limited by only including English articles. Articles in other languages containing important content may have been missed. The review also deviated slightly from the original protocol. For example, the eligibility criteria were updated to exclude case studies and studies with patients under the age of 18 to focus the scope of the review further. These deviations were made before any data extraction occurred; however, the data from paediatric and case studies may have been of value and should be the focus of future reviews.

### Conclusion

This review systematically summarised the relevant research investigating the effectiveness of CBT-based interventions for patients with gastroduodenal DGBIs. Generally, CBT improved gastrointestinal symptoms and psychological outcomes, with benefits persisting at follow-up up to 12-months. This promising evidence suggests that CBT-based interventions may be an effective treatment option for patients with gastroduodenal DGBIs, alongside traditional medical care. CBT could therefore be incorporated into multidisciplinary care and targeted specifically to patients with psychological comorbidities or strong gut-brain influences on their symptoms. More research is needed into the acceptability and effectiveness of digital CBT interventions, which may increase the accessibility of CBT for patients with gastroduodenal DGBIs.

## Data Availability

All data produced in the present study are available upon reasonable request to the authors

## References

[1] Sperber AD, Bangdiwala SI, Drossman DA, Ghoshal UC, Simren M, Tack J, et al. Worldwide prevalence and burden of functional gastrointestinal disorders, results of Rome foundation global study. Gastroenterology 2021;160:99–114. https://doi.org/10.1053/j.gastro.2020.04.014.

[2] Drossman DA. Functional gastrointestinal disorders: History, pathophysiology, clinical features and Rome IV. Gastroenterology 2016;150:1262–79. https://doi.org/10.1053/j.gastro.2016.02.032.

[3] Tack J, Talley NJ, Camilleri M, Holtmann G, Hu P, Malagelada J-R, et al. Functional gastroduodenal disorders. Gastroenterology 2006;130:1466–79. https://doi.org/10.1053/j.gastro.2005.11.059.

[4] Stanghellini V, Chan FKL, Hasler WL, Malagelada JR, Suzuki H, Tack J, et al. Gastroduodenal disorders. Gastroenterology 2016;150:1380–92. https://doi.org/10.1053/j.gastro.2016.02.011.

[5] The Rome Foundation. Appendix A: Rome IV Diagnostic Criteria for FGIDs 2016, January 16. https://theromefoundation.org/rome-iv/rome-iv-criteria/.

[6] Park M-I, Camilleri M. Gastroparesis: Clinical update. Am J Gastroenterol 2006;101:1129–39. https://doi.org/10.1111/j.1572-0241.2006.00640.x.

[7] Huang I-H, Schol J, Khatun R, Carbone F, Van den Houte K, Colomier E, et al. Worldwide prevalence and burden of gastroparesis-like symptoms as defined by the United European Gastroenterology (UEG) and European Society for Neurogastroenterology and Motility (ESNM) consensus on gastroparesis. United European Gastroenterol J 2022. https://doi.org/10.1002/ueg2.12289.

[8] Andreasson A, Talley NJ, Walker MM, Jones MP, Platts LG, Wallner B, et al. An increasing incidence of upper gastrointestinal disorders over 23 years: A prospective population-based study in Sweden. Am J Gastroenterol 2021;116:210–3. https://doi.org/10.14309/ajg.0000000000000972.

[9] Wang YR, Fisher RS, Parkman HP. Gastroparesis-related hospitalizations in the United States: Trends, characteristics, and outcomes, 1995-2004. Am J Gastroenterol 2008;103:313–22. https://doi.org/10.1111/j.1572-0241.2007.01658.x.

[10] Chang L. Review article: Epidemiology and quality of life in functional gastrointestinal disorders. Aliment Pharmacol Ther 2004;20 Suppl 7:31–9. https://doi.org/10.1111/j.1365-2036.2004.02183.x.

[11] Yu D, Ramsey FV, Norton WF, Norton N, Schneck S, Gaetano T, et al. The burdens, concerns, and quality of life of patients with gastroparesis. Dig Dis Sci 2017;62:879–93. https://doi.org/10.1007/s10620-017-4456-7.

[12] Lacy BE, Crowell MD, Mathis C, Bauer D, Heinberg LJ. Gastroparesis: Quality of life and health care utilization. J Clin Gastroenterol 2018;52:20–4. https://doi.org/10.1097/MCG.0000000000000728.

[13] Carabotti M, Scirocco A, Maselli MA, Severi C. The gut-brain axis: Interactions between enteric microbiota, central and enteric nervous systems. Ann Gastroenterol Hepatol 2015;28:203–9.

[14] Weltens N, Iven J, Van Oudenhove L, Kano M. The gut-brain axis in health neuroscience: Implications for functional gastrointestinal disorders and appetite regulation. Ann N Y Acad Sci 2018;1428:129–50. https://doi.org/10.1111/nyas.13969.

[15] Keightley PC, Koloski NA, Talley NJ. Pathways in gut-brain communication: Evidence for distinct gut-to-brain and brain-to-gut syndromes. Aust N Z J Psychiatry 2015;49:207–14. https://doi.org/10.1177/0004867415569801.

[16] Wu JCY. Community-based study on psychological comorbidity in functional gastrointestinal disorder. J Gastroenterol Hepatol 2011;26 Suppl 3:23–6. https://doi.org/10.1111/j.1440-1746.2011.06642.x.

[17] Knowles SR, Apputhurai P, Palsson OS, Bangdiwala S, Sperber AD, Mikocka-Walus A. The epidemiology and psychological comorbidity of disorders of gut-brain interaction in Australia: Results from the Rome Foundation Global Epidemiology Study. Neurogastroenterol Motil 2023:e14594. https://doi.org/10.1111/nmo.14594.

[18] Shoji T, Endo Y, Fukudo S. Psycho-gastroenterology. In: Tominaga K, Kusunoki H, editors. Functional Dyspepsia: Evidences in Pathophysiology and Treatment, Singapore: Springer Singapore; 2018, p. 105–15. https://doi.org/10.1007/978-981-13-1074-4_9.

[19] Luo Y, Keefer L. Role of psychological questionnaires in clinical practice and research within functional gastrointestinal disorders. Neurogastroenterol Motil 2021;33:e14297. https://doi.org/10.1111/nmo.14297.

[20] Van Oudenhove L, Crowell MD, Drossman DA, Halpert AD, Keefer L, Lackner JM, et al. Biopsychosocial aspects of functional gastrointestinal disorders. Gastroenterology 2016;150:1355–67. https://doi.org/10.1053/j.gastro.2016.02.027.

[21] Wouters MM, Boeckxstaens GE. Is there a causal link between psychological disorders and functional gastrointestinal disorders? Expert Rev Gastroenterol Hepatol 2016;10:5–8. https://doi.org/10.1586/17474124.2016.1109446.

[22] Wu JC. Psychological co-morbidity in functional gastrointestinal disorders: Epidemiology, mechanisms and management. J Neurogastroenterol Motil 2012;18:13–8. https://doi.org/10.5056/jnm.2012.18.1.13.

[23] Chang L, Toner BB, Fukudo S, Guthrie E, Locke GR, Norton NJ, et al. Gender, age, society, culture, and the patient’s perspective in the functional gastrointestinal disorders. Gastroenterology 2006;130:1435–46. https://doi.org/10.1053/j.gastro.2005.09.071.

[24] Rodrigues DM, Motomura DI, Tripp DA, Beyak MJ. Are psychological interventions effective in treating functional dyspepsia? A systematic review and meta-analysis. J Gastroenterol Hepatol 2021;36:2047–57. https://doi.org/10.1111/jgh.15566.

[25] Wei Z, Xing X, Tantai X, Xiao C, Yang Q, Jiang X, et al. The effects of psychological interventions on symptoms and psychology of functional dyspepsia: A systematic review and meta-analysis. Front Psychol 2022;13:827220. https://doi.org/10.3389/fpsyg.2022.827220.

[26] Keefer L, Ballou SK, Drossman DA, Ringstrom G, Elsenbruch S, Ljótsson B. A rome working team report on brain-gut behavior therapies for disorders of gut-brain interaction. Gastroenterology 2022;162:300–15. https://doi.org/10.1053/j.gastro.2021.09.015.

[27] Palsson OS, Ballou S. Hypnosis and cognitive behavioral therapies for the management of gastrointestinal disorders. Curr Gastroenterol Rep 2020;22:31. https://doi.org/10.1007/s11894-020-00769-z.

[28] Fenn K, Byrne M. The key principles of cognitive behavioural therapy. InnovAiT 2013;6:579–85. https://doi.org/10.1177/1755738012471029.

[29] Olatunji BO, Hollon SD. Preface: The current status of cognitive behavioral therapy for psychiatric disorders. Psychiatr Clin North Am 2010;33:xiii – xix. https://doi.org/10.1016/j.psc.2010.04.015.

[30] Feingold J, Murray HB, Keefer L. Recent advances in cognitive behavioral therapy for digestive disorders and the role of applied positive psychology across the spectrum of GI care. J Clin Gastroenterol 2019;53:477–85. https://doi.org/10.1097/MCG.0000000000001234.

[31] Li L, Xiong L, Zhang S, Yu Q, Chen M. Cognitive-behavioral therapy for irritable bowel syndrome: A meta-analysis. J Psychosom Res 2014;77:1–12. https://doi.org/10.1016/j.jpsychores.2014.03.006.

[32] Higgins JP, Thomas J, Chandler J, Cumpston M, Li T, Page MJ, et al., editors. Cochrane handbook for systematic reviews of interventions. Cochrane; 2022. https://doi.org/www.training.cochrane.org/handbook.

[33] Page MJ, Moher D, Bossuyt PM, Boutron I, Hoffmann TC, Mulrow CD, et al. PRISMA 2020 explanation and elaboration: Updated guidance and exemplars for reporting systematic reviews. BMJ 2021;372:n160. https://doi.org/10.1136/bmj.n160.

[34] Sterne JAC, Savović J, Page MJ, Elbers RG, Blencowe NS, Boutron I, et al. RoB 2: A revised tool for assessing risk of bias in randomised trials. BMJ 2019;366:l4898. https://doi.org/10.1136/bmj.l4898.

[35] Sterne JA, Hernán MA, Reeves BC, Savović J, Berkman ND, Viswanathan M, et al. ROBINS-I: A tool for assessing risk of bias in non-randomised studies of interventions. BMJ 2016;355:i4919. https://doi.org/10.1136/bmj.i4919.

[36] Glasinovic E, Wynter E, Arguero J, Ooi J, Nakagawa K, Yazaki E, et al. Treatment of supragastric belching with cognitive behavioral therapy improves quality of life and reduces acid gastroesophageal reflux. American Journal of Gastroenterology 2018;113:539–47. https://doi.org/10.1038/ajg.2018.15.

[37] Sawada A, Anastasi N, Green A, Glasinovic E, Wynter E, Albusoda A, et al. Management of supragastric belching with cognitive behavioural therapy: Factors determining success and follow-up outcomes at 6-12 months post-therapy. Aliment Pharmacol Ther 2019;50:530–7. https://doi.org/10.1111/apt.15417.

[38] Dast SPN, Maghsoud FM, Assareh M, Khorramdel K, Rajabi S. Efficacy of mindfulness-based therapy efficacy in reducing physical symptoms and increasing specific quality of life in patients with functional dyspepsia. Bullet Environ Pharmacol Life Sci 2015;3:46– 51.

[39] Punkkinen J, Nyyssönen M, Walamies M, Roine R, Sintonen H, Koskenpato J, et al. Behavioral therapy is superior to follow-up without intervention in patients with supragastric belching-A randomized study. Neurogastroenterol Motil 2022;34:e14171. https://doi.org/10.1111/nmo.14171.

[40] Niesen CR, Olson DM, Nowdesha KD, Tynsky DA, Loftus CG, Meiers SJ. Enhancing self-management for adults with functional abdominal pain: A registered nurse-led cognitive-behavioral therapy approach. Gastroenterol Nurs 2018;41:321–32. https://doi.org/10.1097/SGA.0000000000000322.

[41] Orive M, Barrio I, Orive VM, Matellanes B, Padierna JA, Cabriada J, et al. A randomized controlled trial of a 10 week group psychotherapeutic treatment added to standard medical treatment in patients with functional dyspepsia. J Psychosom Res 2015;78:563–8. https://doi.org/10.1016/j.jpsychores.2015.03.003.

[42] Haug TT, Wilhelmsen I, Svebak S, Berstad A, Ursin H. Psychotherapy in functional dyspepsia. J Psychosom Res 1994;38:735–44. https://doi.org/10.1016/0022-3999(94)90026-4.

[43] Haag S, Senf W, Tagay S, Langkafel M, Braun-Lang U, Pietsch A, et al. Is there a benefit from intensified medical and psychological interventions in patients with functional dyspepsia not responding to conventional therapy? Aliment Pharmacol Ther 2007;25:973–86. https://doi.org/10.1111/j.1365-2036.2007.03277.x.

[44] Batebi S, Masjedi Arani A, Jafari M, Sadeghi A, Saberi Isfeedvajani M, Davazdah Emami MH. A randomized clinical trial of metacognitive therapy and nortriptyline for anxiety, depression, and difficulties in emotion regulation of patients with functional dyspepsia. Res Psychother 2020;23:157–66. https://doi.org/10.4081/ripppo.2020.448.

[45] Dehghanizade Z, Zargar Y, Mehrabizadeh Honarmand M, Kadkhodaie A, Eydi Baygi M. The effectiveness of cognitive behavior stress management on functional dyspepsia symptoms. J Adv Med Educ Prof 2015;3:45–9.

[46] Tavakoli T, Hoseini M, Tabatabaee TSJ, Rostami Z, Mollaei H, Bahrami A, et al. Comparison of dialectical behavior therapy and anti-anxiety medication on anxiety and digestive symptoms in patients with functional dyspepsia. J Res Med Sci 2020;25:59. https://doi.org/10.4103/jrms.JRMS_673_19.

[47] Murray HB, Zhang F, Call CC, Keshishian A, Hunt RA, Juarascio AS, et al. Comprehensive cognitive-behavioral interventions augment diaphragmatic breathing for rumination syndrome: A proof-of-concept trial. Dig Dis Sci 2021;66:3461–9. https://doi.org/10.1007/s10620-020-06685-6.

[48] Teh KK-J, Ng Y-K, Doshi K, Tay S-W, Hao Y, Ang L-Y, et al. Mindfulness-based cognitive therapy in functional dyspepsia: A pilot randomized trial. J Gastroenterol Hepatol 2021;36:2058–66. https://doi.org/10.1111/jgh.15389.

[49] McGuinness LA, Higgins JPT. Risk-of-bias VISualization (robvis): An R package and Shiny web app for visualizing risk-of-bias assessments. Research Synthesis Methods 2020;n/a. https://doi.org/10.1002/jrsm.1411.

[50] Tang Q-L, Lin G-Y, Zhang M-Q. Cognitive-behavioral therapy for the management of irritable bowel syndrome. World J Gastroenterol 2013;19:8605–10. https://doi.org/10.3748/wjg.v19.i46.8605.

[51] Kinsinger SW. Cognitive-behavioral therapy for patients with irritable bowel syndrome: Current insights. Psychol Res Behav Manag 2017;10:231–7. https://doi.org/10.2147/PRBM.S120817.

[52] Sugaya N, Shirotsuki K, Nakao M. Cognitive behavioral treatment for irritable bowel syndrome: A recent literature review. Biopsychosoc Med 2021;15:23. https://doi.org/10.1186/s13030-021-00226-x.

[53] Black CJ, Paine PA, Agrawal A, Aziz I, Eugenicos MP, Houghton LA, et al. British Society of Gastroenterology guidelines on the management of functional dyspepsia. Gut 2022. https://doi.org/10.1136/gutjnl-2022-327737.

[54] Law M, Bartlett E, Sebaratnam G, Pickering I, Simpson K, Keane C, et al. “One more tool in the tool belt”: A qualitative interview study investigating patient and clinician opinions on the integration of psychometrics into routine testing for disorders of gut-brain interaction. medRxiv 2023. https://doi.org/10.1101/2023.06.06.23291063.

[55] Gharibans AA, Calder S, Varghese C, Waite S, Schamberg G, Daker C, et al. Gastric dysfunction in patients with chronic nausea and vomiting syndromes defined by a noninvasive gastric mapping device. Science Translational Medicine 2022;14:eabq3544. https://doi.org/10.1126/scitranslmed.abq3544.

[56] Bray NA, Koloski NA, Jones MP, Do A, Pang S, Coombes JS, et al. Evaluation of a multidisciplinary integrated treatment approach versus standard model of care for functional gastrointestinal disorders (FGIDS): A matched cohort study. Dig Dis Sci 2022;67:5593–601. https://doi.org/10.1007/s10620-022-07464-1.

[57] Basnayake C, Kamm MA, Stanley A, Wilson-O’Brien A, Burrell K, Lees-Trinca I, et al. Standard gastroenterologist versus multidisciplinary treatment for functional gastrointestinal disorders (MANTRA): an open-label, single-centre, randomised controlled trial. Lancet Gastroenterol Hepatol 2020;5:890–9. https://doi.org/10.1016/S2468-1253(20)30215-6.

[58] Basnayake C, Kamm MA, Stanley A, Wilson-O’Brien A, Burrell K, Lees-Trinca I, et al. Long-term outcome of multidisciplinary versus standard gastroenterologist care for functional gastrointestinal disorders: A randomized trial. Clin Gastroenterol Hepatol 2022;20:2102–11.e9. https://doi.org/10.1016/j.cgh.2021.12.005.

[59] Basnayake C, Kamm MA, Salzberg MR, Wilson-O’Brien A, Stanley A, Thompson AJ. Delivery of care for functional gastrointestinal disorders: A systematic review. J Gastroenterol Hepatol 2020;35:204–10. https://doi.org/10.1111/jgh.14830.

[60] Kim H, Oh Y, Chang SJ. Internet-Delivered Cognitive Behavioral Therapy in Patients With Irritable Bowel Syndrome: Systematic Review and Meta-Analysis. J Med Internet Res 2022;24:e35260. https://doi.org/10.2196/35260.

[61] Hunt M, Miguez S, Dukas B, Onwude O, White S. Efficacy of Zemedy, a mobile digital therapeutic for the self-management of irritable bowel syndrome: Crossover randomized controlled trial. JMIR Mhealth Uhealth 2021;9:e26152. https://doi.org/10.2196/26152.

[62] Szigethy E, Tansel A, Pavlick AN, Marroquin MA, Serio CD, Silfee V, et al. A coached digital cognitive behavioral intervention reduces anxiety and depression in adults with functional gastrointestinal disorders. Clin Transl Gastroenterol 2021;12:e00436. https://doi.org/10.14309/ctg.0000000000000436.

[63] Laird KT, Tanner-Smith EE, Russell AC, Hollon SD, Walker LS. Short-term and long-term efficacy of psychological therapies for irritable bowel syndrome: A systematic review and meta-analysis. Clin Gastroenterol Hepatol 2016;14:937–47.e4. https://doi.org/10.1016/j.cgh.2015.11.020.

[64] Chen LJ, Kamp K, Fang A, Heitkemper MM. Delivery methods of cognitive behavior therapy for patients with irritable bowel syndrome. Gastroenterol Nurs 2022;45:149–58. https://doi.org/10.1097/SGA.0000000000000671.

